# Cerebellar tissue properties alterations in fibromyalgia: a T1w/T2w ratio study

**DOI:** 10.1101/2025.04.17.25326040

**Authors:** Yann Quidé, Sylvia M. Gustin

**Author notes:** Corresponding author: Dr Yann Quidé, NeuroRecovery Research Hub, School of Psychology, Biological Sciences (Biolink) building, Level 1, UNSW Sydney, NSW, 2052, Australia.

## Abstract

Fibromyalgia is associated with elevated levels of comorbid anxiety and depression, together impacting brain morphology possibly reflecting common underlying biological processes. The present study aims to determine the difference in regional myelination in females with fibromyalgia compared to females who do not experience chronic pain and determine the role of the severity of comorbid anxiety and depressive symptoms experienced to mediate this difference in brain myelination. Thirty-three females with and 33 females without (Controls) fibromyalgia were included, for which the severity of depressive and anxiety symptoms were recorded using the Hamilton Anxiety/Depression Rating scales (HAMA/HAMD). Whole-brain three-dimensional T1-weighted (T1w) and T2-weighted (T2w) magnetic resonance imaging scans were collected, and T1w/T2w ratio (myelin maps) derived. Mediation analyses were performed with anxiety and depressive symptoms as mediators of the T1w/T2w ratio differences among the groups. Compared to the control group, the fibromyalgia group lower T1w/T2w values in the left cerebellar lobule VI (*pFWEc*=0.030) and left cerebellar lobule VIII (*pFWEc*=0.029). These T1w/T2w values were significantly negatively associated with severity of anxiety and depressive symptoms (all *p*<0.001). Mediation analyses indicated that the severity of anxiety (but not depressive) symptoms mediated the group difference in T1w/T2w values in cerebellar lobule VI (*p*=0.012), but not VIII (*p*=0.813). Lowered cerebellar myelination may reflect chronic states of low-grade inflammation, resulting from the long-term consequences of living with fibromyalgia and related anxiety and depressive symptoms. This remains speculative, and future studies integrating peripheral biological markers of inflammation are warranted to confirm this interpretation.

## Introduction

Fibromyalgia is characterised by widespread pain for over three months, that can arise in the absence of an evident organic lesion or injury [56]. Fibromyalgia can affect up to 5% of the general population, predominantly females, and is associated with poor quality of life [58]. As most chronic pain conditions, fibromyalgia is also associated with higher rates of comorbid conditions, including mental health problems, such as pathological levels of anxiety (estimated between 20%–80%) and depression (estimated between 13%–64%) [19; 51]. Anxiety and depression are both risk factors and consequences of living with fibromyalgia [3], and together may impact similar systems, including brain integrity.

Fibromyalgia is associated with smaller thalamic or cerebellar volumes [10; 13; 46]. Some of these brain alterations are accounted for by comorbid mental health conditions in fibromyalgia. Increasing severity of depressive symptoms is associated with smaller left putamen [40], larger cerebellar, medial prefrontal/orbitofrontal cortices, medial temporal lobe, and right inferior parietal lobule volumes [32]. On the other hand, the severity of anxiety symptoms is associated with increasing grey matter volume in the medial orbitofrontal cortex [16] and the amygdala [29]. Overall, these findings suggest that brain morphology alterations may results from neurobiological mechanisms common to fibromyalgia and its comorbid conditions.

Aberrant increased low-grade inflammation is one of these common neurobiological mechanisms. Low-grade inflammation refers to the (low-grade) continuous production of inflammatory markers, not in response to an infection [20], and commonly reported across depressive, anxious and chronic pain conditions [4; 38; 54]. Low-grade inflammation activates glial cells (astrocytes, microglia, and oligodendrocytes) that can damage brain integrity and alter the formation of myelin sheaths in the brain [14; 42]. However, to the best of our knowledge, there is to date no study investigating the potential alterations in brain myelination in fibromyalgia, or their associations with anxiety and depression.

Neuroimaging methodologies can produce proxy measures of cortical microstructural properties, including myelination. For instance, “myelin map” can be derived from routine T1-weighted (T1w) and T2-weighted (T2w) magnetic resonance imaging (MRI) scans, by calculating the ratio of T1w to T2w signal intensities (T1w/T2w ratio) [21–24]. This relatively simple approach provides new opportunities to better understand brain alterations in complex conditions such as fibromyalgia.

The present study aims to determine the difference in regional myelination in females with fibromyalgia compared to females who do not experience chronic pain and determine the role of the severity of comorbid anxiety and depressive symptoms experienced to mediate this difference in brain myelination. As there is no other study investigating myelination in fibromyalgia to our knowledge, we used a data-driven, whole-brain approach. Fibromyalgia was expected to be associated with lower local levels of myelination in brain regions commonly reporting smaller grey matter volume and associated with anxiety and depressive symptoms. These regions include the cerebellum, medial prefrontal/orbitofrontal cortex, medial temporal lobe, inferior parietal lobule, and amygdala. In addition, severity of anxiety and depressive symptoms were expected to account at least partially for (mediate) these group-related myelination alterations.

## Material and Methods

### Participants

Participants from this cross-sectional study were 33 females with fibromyalgia and 33 females without a chronic pain condition (Controls) from the OpenNeuro dataset ds004144 [6]. Details about recruitment and clinical assessments for this dataset are available elsewhere [5]. Briefly, participants included were between 18 and 50 years old who completed at least elementary school. All participants with fibromyalgia received their diagnosis from a rheumatologist or an internal medicine specialist, confirmed by the American College of Rheumatology 1990 [57] and 2016 criteria [55]. These criteria were also used to exclude controls. Exclusion criteria included the presence of a major psychiatric, cardiovascular, neurological or other pain conditions. All participants answered the Socio-economic levels Questionnaire of the Mexican Association of Market Research and Public Opinion Agencies Questionnaire (AMAI NSE 8×7), which is a standardized instrument to measure socio-economic level in Mexican households. Socio-demographic details are presented Table 1. All participants provided written informed consent. The protocol was approved by the Research Ethics Committee of the National Institute of Psychiatry “Ramón de la Fuente Muñiz” in Mexico City, in accordance with the Declaration of Helsinki.

### Clinical scales

Participants completed a comprehensive clinical and psychological assessment no longer than two weeks prior their magnetic resonance imaging (MRI) session. The total score to the 20-item Spanish version of the Fibromyalgia Impact Questionnaire (FIQ) was used to evaluate the current health status and symptoms severity associated with fibromyalgia [53]. The 78-item Spanish version of the McGill pain questionnaire was used evaluate the pain from a dimensional perspective [36]: affective-emotional, evaluative (general description), sensory (temporo-spatial qualities), and miscellaneous. The Fibromyalgia General Questionnaire was developed to record about the duration and current treatment for fibromyalgia. Three open-ended questions and one close-ended question assess the duration of fibromyalgia symptoms, the time with the diagnosis, and the current medication received.

### Other clinical scales

The original study collected more clinical measures, but the present study focused on indices of depressive and anxiety symptoms. Severity of depressive symptoms were evaluated using the Spanish version of the Hamilton Depression Rating Scale (HAMD) [12]. The version used comprised 21 items, each item being scored on a Likert-type scale between 0 and 4, the higher the total score, the more severe depressive symptoms. The Spanish version of the Hamilton Anxiety Rating Scale (HAMA) [9] was used to measure the severity of anxiety symptoms experienced. This 14-item Likert-type scale scored the intensity, frequency and dysfunction caused by symptoms, ranging from 0 to 4. The higher the total score, the more intense the symptoms.

### Magnetic resonance imaging

#### Images acquisition

Whole-brain T1-weighted and T2-weighted scans were acquired using a Philips 3T Ingenia scanner (Philips Healthcare, Best, The Netherlands) with a 32- channel phased array coil. As described in [5; 6], scanning parameters for the three-dimensional T1-weighted FFE SENSE sequence were: repetition time (TR) = 7.0 ms, echo time (TE) = 3.5 ms, flip angleLJ=LJ8°, field of view (FOV)LJ=LJ240LJmm2, matrixLJ=LJ240LJ×LJ240LJmm, number of slicesLJ=LJ180, gapLJ=LJ0, planeLJ=LJsagittal, slice thicknessLJ=LJ1.0 mm, voxel sizeLJ=LJ1LJ×LJ1LJ×LJ1LJmm.

Scanning parameters for the three-dimensional T2-weighted FFE SENSE sequence were: TR = 2.5 ms, TE= 0.3 ms, flip angleLJ=LJ90°, FOVLJ=LJ240LJmm2, matrixLJ=LJ240LJ×LJ240LJmm, number of slicesLJ=LJ180, gapLJ=LJ0, planeLJ=LJsagittal, slice thicknessLJ=LJ1.0 mm, voxel sizeLJ=LJ1LJ×LJ1LJ×LJ1LJmm.

#### Images preprocessing

Both T1w and T2w images for each participants were processed using the MRTool toolbox (version 1.4.3; https://www.nitrc.org/projects/mrtool/) [21; 22] for SPM12 (v7771; Wellcome Department of Cognitive Neurology, University College London, UK; https://www.fil.ion.ucl.ac.uk/spm/) in Matlab r2024a (MathWorks Inc., Sherborn, Massachusetts, USA). The default *MRTool-T1/T2* pipeline was used. First, each participant’s T2w image was co-registered to their respective T1w image, using a rigid-body transformation. Then, to ensure spatial equalization of the coil sensitivity profiles, both T1w and T2w images were subjected to bias correction for intensity non-uniformity (default input parameters: 60mm full width at half maximum (FWHM) smoothing and very light regularisation (0.0001)). This step removed the slow intensity variations related not only to the MR hardware but also its interaction with the subject’s cranial tissue. The intensity of both bias-corrected images were separately standardized using a non-linear external calibration approach (MRTool image calibration option #1: Non-linear histogram matching-external calibration), in order to accurately capture inter-individual differences in myelin contrast [21; 22].

The T1-w/T2-w image was calculated as the ratio of the standardized T1w and T2w images. These whole-brain contrast images were then normalised to The Montreal Neurological Institute (MNI) space and skull-stripped to remove non-brain tissue.

### Statistical analyses

Due to the relatively small sample size, group differences in whole-brain T1w/T2w ratio were analysed using two-sample t-tests with the Statistical non-Parametric Mapping toolbox (SnPM13.1.09; http://www.nisox.org/Software/SnPM13/) for SPM12 [41]. This toolbox uses permutation tests that do not rely on assumptions of normality and are therefore less likely to produce false positive results, especially for smaller samples [17; 59]. Within the SnPM13 toolbox, age was added as a covariate, default parameters were used to define the cluster-forming threshold, and family-wise error correction was applied to the cluster statistics (*pFWEc* < 0.05). Cluster-based parameters were set to perform 10,000 permutations, and variance smoothing was not applied (set to [0,0,0]). Following whole-brain analyses, raw signal at the peak of significant clusters were extracted for further analyses using R (version 4.4.0) [45] in RStudio (version 2024.04.1+748) [44], including regression analyses to determine the relationship between T1w/T2w ratio values and severity of anxiety and depressive symptoms.

To determine the role of depressive and anxiety symptoms in brain myelination (T1w/T2w ratio) group differences, mediation analyses were conducted on the signal from peaks of significant whole-brain clusters using the *lavaan* package (version 0.6-18) [47]. This package produces unbiased estimates of indirect effects and controls for variable inter-dependencies. Group was the independent variable, T1w/T2w ratio was the dependent variable, and depressive (HAMD total score) or anxiety symptoms (HAMA total score) were the mediators; age was used as a covariate of non-interest. Standardized estimates for all variables (std.all) and 95% confidence intervals (95%bootCI) were determined using bootstrapping (10,000 bootstraps) using the *bca*.*simple* function in *lavaan*. For sake of completeness, mediation analyses using T1w/T2w ratio values as mediators were performed (see Supplementary Table S1 and Table S2).

Finally, exploratory whole-brain regression analyses were conducted using SnPM13 (10,000 permutations, variance smoothing [0,0,0]) to determine the potential associations between symptom severity (FIQ total score), symptoms duration (in years), and medication use (0 = no pharmacological medication or use only during crisis; 1 = daily pharmacological medication and additional use during crisis) on brain myelination within the fibromyalgia group only. In keeping with the above whole-brain analyses, age was included as a covariate of no interest, and statistical significance was set at *pFWEc* < 0.05 for these analyses.

## Results

### Sample characteristics

Details of the socio-demographic and clinical characteristics of the sample are presented Table 1 and Table 2, respectively. Briefly, the fibromyalgia and the control groups were statistically matched for age, handedness, years of education, occupation levels, socio-economical levels and probable menstrual cycles (see Table 1). Compared to the control group, the fibromyalgia group showed higher levels of depressive and anxiety symptoms.

### Whole-brain T1w/T2w ratio analyses

Whole-brain non-parametric analyses showed reduced T1w/T2w ratio in the left cerebellar lobule VI (peak MNI coordinates [-33,-35,-33], k = 525 voxels, *t(63)* = 5.34, *pFWEc* = 0.030) and left cerebellar lobule VIII (peak MNI coordinates [-40,-41,- 46], k = 551 voxels, *t(63)* = 5.01, *pFWEc* = 0.029) in the fibromyalgia group, compared to the control group (see Figure 1).

**Figure 1.**
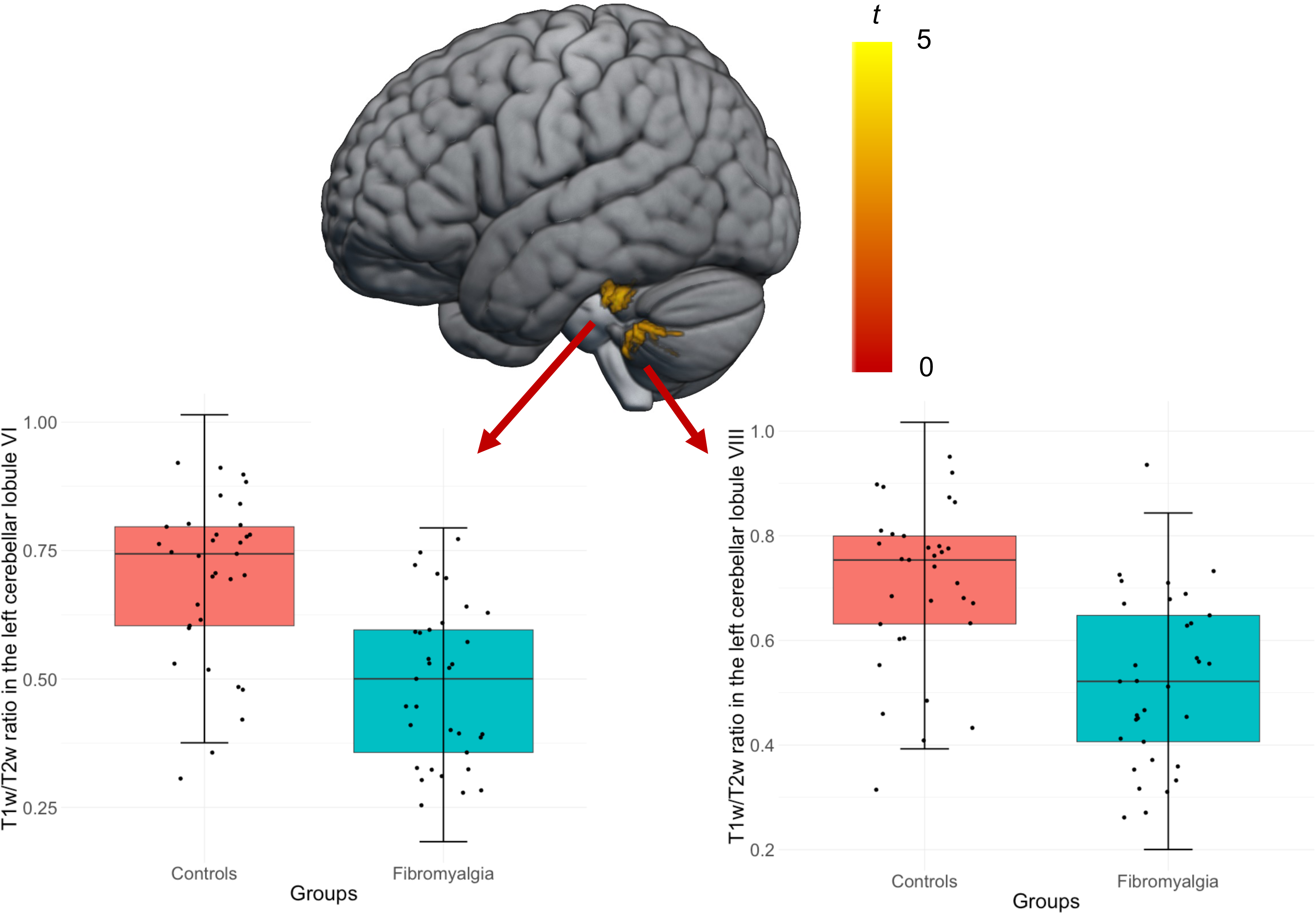
**Whole-brain T1w/T2w ratio maps differences between females with fibromyalgia and controls** Accounting for age, females with fibromyalgia (in green) showed significantly (*pFWEc* < 0.05) lower T1w/T2w ratio values in the left cerebellar lobules VI (left panel) and VIII (right panel) compared to the control group (in red). The colour-bar represents *t*-statistics, the mean T1w/T2w ratio values are represented with horizontal segment within the boxplot, the bars represent standard deviations to the mean, and individual data points are represented.

### Associations between whole-brain T1w/T2w ratio values and mental health symptoms

T1w/T2w ratio values in the left cerebellar lobules VI and VIII were significantly negatively associated with severity of anxiety (lobule VI: *b* = −0.010, *p* < 0.001; lobule VIII: *b* = −0.007, *p* < 0.001) and depressive symptoms (lobule VI: *b* = - 0.010, *p* < 0.001; lobule VIII: *b* = −0.008, *p* < 0.001).

### Mediation analyses

Statistical details from the mediation analyses are provided in Table 3 and Table 4 and illustrated in Figure 2. Statistical significance was adjusted to account for the number of models tested using Bonferroni correction (*p*=0.05/4=0.0125).

**Figure 2.**
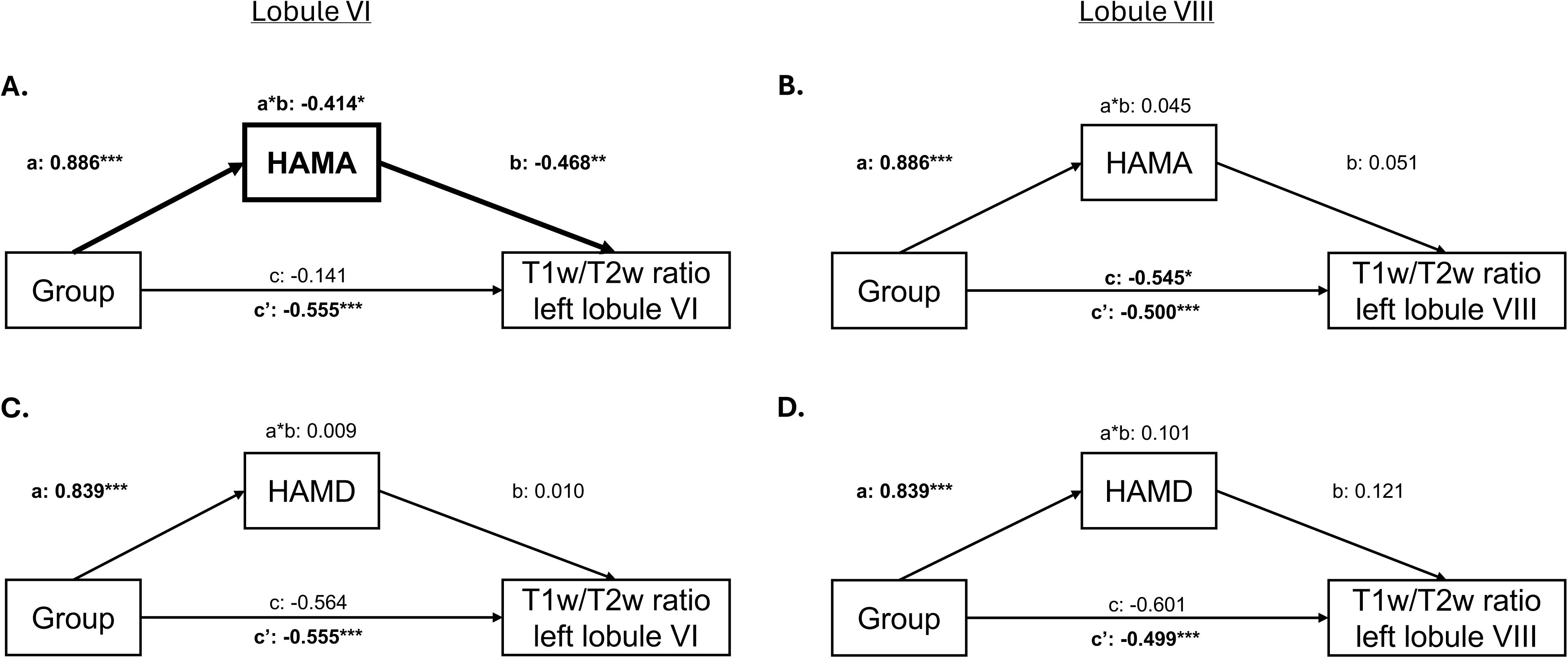
**Mediation analyses** On the top panel, mediation effects of the severity of anxiety symptoms (HAMA total score) on group differences in myelination of the left cerebellar (A) lobule VI or (B) lobule VIII. On the bottom panel, mediation effects of the severity of depressive symptoms (HAMD total score) on group differences in myelination of the left cerebellar (C) lobule VI or (D) lobule VIII. \**p* < 0.05; \*\**p* < 0.01, \*\*\**p* < 0.001

*Anxiety.* The model investigating the role of anxiety to mediate the group difference in T1w/T2w ratio in the left cerebellar lobule VI showed that group was significantly associated with anxiety symptoms (path a: *standardized b* = 0.886, *p* < 0.001), indicating more severe anxiety symptoms in people with fibromyalgia compared with controls. Then, severity of anxiety symptoms (path b: *standardized b* = −0.468, *p* = 0.009) and group (total effect) were significantly associated with T1w/T2w ratio in the left cerebellar lobule VI (path c’: *standardized b* = −0.141, *p* < 0.001), indicating that the more severe anxiety symptoms and having fibromyalgia were associated with less myelination in cerebellar lobule VI. The indirect effect of direct effect of group on T1w/T2w ratio in the left cerebellar lobule VI was significant (a*b: *standardized b* = −0.414, *p* = 0.012), but the direct effect was no longer significant once levels of anxiety were removed from the model (path c: *standardized b* = −0.141, *p* = 0.442), indicating full mediation (see Figure 2A).

Similarly to above, the model investigating the role of anxiety to mediate the group difference in T1w/T2w ratio in the left cerebellar lobule VIII showed that group was significantly associated with anxiety symptoms (path a: *standardized b* = 0.886, *p* < 0.001), indicating more severe anxiety symptoms in people with fibromyalgia compared with controls. Severity of anxiety symptoms was not (path b: *standardized b* = 0.051, *p* = 0.810) but group (total effect) was significantly associated with T1w/T2w ratio in the left cerebellar lobule VIII (path c’: *standardized b* = −0.500, *p* < 0.001), indicating that the more severe anxiety symptoms and having fibromyalgia were associated with less myelination in cerebellar lobule VIII. The indirect effect of direct effect of group on T1w/T2w ratio in the left cerebellar lobule VIII was not significant (a*b: *standardized b* = 0.045, *p* = 0.813), and the direct effect was longer significant (but did not survive Bonferroni correction) once levels of anxiety were removed from the model (path c: *standardized b* = −0.545, *p* = 0.015; see Figure 2B).

*Depression.* The model investigating the role of depression to mediate the group difference in T1w/T2w ratio in the left cerebellar lobule VI also showed that group was significantly associated with depressive symptoms (path a: *standardized b* = 0.839, *p* < 0.001), indicating more severe depressive symptoms in people with fibromyalgia compared with controls. Severity of depressive symptoms were not associated with T1w/T2w ratio in the left cerebellar lobule VI (path b: *standardized b* = 0.010, *p* = 0.952) but group (total effect) was significantly associated with T1w/T2w ratio in the left cerebellar lobule VI (path c’: *standardized b* = −0.555, *p* < 0.001), indicating that having fibromyalgia were associated with less myelination in cerebellar lobule VI, but not the severity of depressive symptoms. The indirect effect of group on T1w/T2w ratio in the left cerebellar lobule VI was not significant (a*b: *standardized b* = 0.009, *p* = 0.953), but the direct effect was significant once levels of depression were removed from the model (path c: *standardized b* = −0.564, *p* < 0.001; see Figure 2C).

The model investigating the role of depression to mediate the group difference in T1w/T2w ratio in the left cerebellar lobule VIII also showed that group was significantly associated with depressive symptoms (path a: *standardized b* = 0.839, *p* < 0.001), indicating more severe depressive symptoms in people with fibromyalgia compared with controls. Severity of depressive symptoms were not associated with

T1w/T2w ratio in the left cerebellar lobule VIII (path b: *standardized b* = 0.121, *p* = 0.500) but group (total effect) was significantly associated with T1w/T2w ratio in the left cerebellar lobule VIII (path c’: *standardized b* = −0.500, *p* < 0.001), indicating that having fibromyalgia was associated with less myelination in cerebellar lobule VIII, but not the severity of depressive symptoms. The indirect effect of group on T1w/T2w ratio in the left cerebellar lobule VIII was not significant (a*b: *standardized b* = 0.101, *p* = 0.513), but the direct effect was significant once levels of depression were removed from the model (path c: *standardized b* = −0.601, *p* = 0.002; see Figure 2D).

### Exploratory analyses

Whole-brain regression analyses showed no significant association between FIQ total score, symptoms duration or medication use and brain myelination.

Additional regression analyses specific to lobules VI and VIII clusters were conducted in R, and reported the same lack of association with FIQ total score (lobule VI: *b* = −0.002, *p* = 0.418; lobule VIII: *b* = −0.001, *p* = 0.801), symptoms duration (lobule VI: *b* = −0.003, *p* = 0.195; lobule VIII: *b* = −0.003, *p* = 0.265) or medication use (lobule VI: *b* = −0.036, *p* = 0.521; lobule VIII: *b* = −0.043, *p* = 0.436).

## Discussion

This study is the first, to our knowledge, to identify alterations in brain tissue properties in females with fibromyalgia. Compared to females not experiencing chronic pain, those with fibromyalgia showed reduced T1w/T2w ratio values, a proxy marker of myelination, in the left cerebellar lobules VI and VIII. Group difference in myelination of the lobule VI, but not lobule VIII, was further mediated by the severity of reported anxiety symptoms. Depressive symptoms did not mediate the group differences in myelination of cerebellar lobules VI and VIII.

Lower myelination of cerebellar lobules VI and VIII were evident in fibromyalgia and were associated with both anxiety and depressive symptoms. This result is partly consistent with our expectations of lower levels of myelination in brain regions showing lower grey matter volume in fibromyalgia with comorbid anxiety and depression, including the cerebellum [32; 46]. Lower cerebellar myelination may represent a consequence of living with fibromyalgia and its comorbid conditions (in average eight years here), similar to living in a state of chronic stress. Indeed, dysfunctions of the hypothalamic_-_pituitary_-_adrenal (HPA) axis largely overlap among chronic stress and chronic pain [1; 30]. Long-term exposure to chronic stress leads to blunted cortisol levels [34]. Although a normal stress response has anti-inflammatory properties, blunted cortisol levels are associated to sustained elevated low-grade, systemic inflammation [7; 15; 18]. Elevated low-grade systemic inflammation can damage brain integrity [35], in particular the formation of myelin sheaths [14; 42]. Interestingly, lower cerebellar myelination has been associated with faster cognitive decline in cognitively unimpaired adults [26], potentially reflecting atypical brain aging, as previously observed in chronic back pain [2]. Although plausible, this interpretation remains speculative, as this study did not record biological markers of low-grade inflammation, cognition or estimated changes in brain ageing. Nevertheless, this study suggests further inquiries into the interplay between chronic stress and chronic pain, through HPA axis dysfunction and sustained low-grade systemic inflammation, and their impacts on cognition so that both preventative and treatment therapeutics can be developed. This is particularly relevant to fibromyalgia as this condition is strongly associated with cognitive and emotional conditions such as anxiety and depression [19; 51]. Further, replication and extension of these findings to other chronic pain conditions are also needed to better understand if these proposed mechanisms also apply across chronic pain conditions or are specific to fibromyalgia.

Not only critical to motor functions, the cerebellum also contributes to cognitive and affective processes [31; 49]. The cerebellum, especially lobule VI, is closely associated with the salience network [28; 48], with preferential connections with the amygdala [27], and critical role for emotional processing [27; 48; 50]. This relationship with the salience network is also consistent with the recent Fibromyalgia: Imbalance of Threat and Soothing Systems model (FITSS) [43]. Within this model, the salience network, along with the management of threat and soothing processes, are core to the development and maintenance of fibromyalgia [43]. Alterations in these processes and associated cerebral and cerebellar regions may, at least in part, explain the heterogeneity of phenotypes observed in fibromyalgia, especially relative to variations in the severity of comorbid conditions experienced in fibromyalgia, such as anxiety. This hypothesis will need to be directly tested in future studies.

Although T1w/T2w ratio indices cannot infer on functional alterations, cerebellar myelination reduction may account for, or at least contribute to, functional alterations commonly observed in fibromyalgia [11]. Reduced myelination of cerebellar lobule VI may impact the strength of the cerebellar-amygdala relationship, altering the normal functioning of the salience network, and leading to reduced stability across large-scale brain networks [37]. Interestingly, similar alterations in the balance between networks is evident in separate studies of comorbid pathologies to fibromyalgia, including anxiety disorders [33]. Future functional brain imaging studies are necessary to understand the impact of reduced cerebellar myelination on brain function in fibromyalgia and comorbid affective conditions.

This study has several limitations. First, the relatively small sample size may have hindered the discovery of smaller critical effects. Larger studies are necessary to better understand the neurobiological mechanisms involved in fibromyalgia and its comorbid conditions. Second, although recorded, the potential effects of medication types and dosages were not accounted for in the statistical analyses, owing to the presence of the control group. Although the use of medication was not associated with T1w/T2w ratio values, it remains unclear if specific drugs and their dosages may influence these indices. Third, the present analyses were cross-sectional and cannot infer about causality or sequence of events. Longitudinal studies following people with fibromyalgia from the early to later stages of the condition would provide this information. Fourth, using T1w/T2w ratio has itself limitations, as it remains only a proxy and not a direct measure of myelination. In addition, the biological processes behind remain unclear, with T1w/T2w ratio remaining a measure sensitive to other features, including dendrite density and axonal diameter (e.g., [52]), as well as changes in iron levels or water concentration (e.g., [39]). Finally, this study included only females, and cannot be generalised to males with fibromyalgia, as sexual dimorphism influences the generation and presentation of chronic pain [8]. This is even more critical when studying fibromyalgia in the context of chronic stress [25].

In conclusion, reduced myelination in left cerebellar lobules VI and VIII was evident in adult females with fibromyalgia. Reduced myelination of left lobule VI was mediated by the severity of anxiety symptoms experienced. These results provide preliminary evidence for cortical microstructural changes in fibromyalgia, potentially resulting from alterations of myelin sheaths formation following chronic exposure to elevated low-grade inflammation. Future studies integrating biological (e.g., inflammatory markers), clinical and functional neuroimaging data are warranted to better understand the impacts of these biological processes on the neurobiological processes underlying fibromyalgia and associated comorbid mental health problems. This information will be critical for the choice of existing, or the development of new interventions, targeting anxiety symptoms that could potentially normalise the underlying neurobiological changes observed here.

## Data Availability

The data that support the findings are fully available from Openneuro.org (https://openneuro.org/datasets/ds004144/versions/1.0.2)

## Acknowledgments

The authors would like to acknowledge the use of the ds004144 dataset, which is publicly accessible through Openneuro.org (doi:10.18112/openneuro.ds004144.v1.0.2) and described in Balducci et al., 2022 (doi: 10.1038/s41597022-01677-9)

## Data availability

The data that support the findings are fully available from Openneuro.org (doi:10.18112/openneuro.ds004144.v1.0.2)

## Authors contributions

Y.Q. contributed conceptualization, data curation, formal analysis, investigation, methodology, validation, visualization, writing of the original draft, and review and editing. S.M.G. contributed conceptualization, data curation, formal analysis, investigation, methodology, project administration, resources, supervision, and review and editing.

