## Supplementary material for "Cerebellar tissue properties alterations in fibromyalgia: a T1w/T2w ratio study": Table 1

| Table 1. Socio-demographic characteristics | | | | | | | |
| --- | --- | --- | --- | --- | --- | --- | --- |
|  | Controls (n=33) | | Fibromyalgia (N=33) | | Statistics | | |
|  | Mean | SD | Mean | SD | t-value/X^2^ | df | p-value |
| Age, in years | 41.52 | 6.03 | 41.73 | 6.09 | 0.142 | 64 | 0.887 |
| Education, in years | 16.55 | 3.95 | 15.52 | 3.96 | -1.058 | 64 | 0.294 |
| Marital status, S/M/SE/W | 7/21/4/1 | | 9/17/5/2 | | 1.115 | 3 | 0.773 |
| Occupation, PE/BU/T/L/H/ST/P/UN/Other | 17/3/2/2/6/1/2/0 | | 10/3/2/2/12/3/0/1 | | 7.815 | 7 | 0.349 |
| Occupation pattern^#^, FT/HTF/HTI | 16/5/6 | | 11/1/11 | | 6.063 | 3 | 0.109 |
| Socioeconomic level^$^, D/D+/C-/C/C+/AB | 1/1/5/1/15/10 | | 0/7/7/1/7/11 | | 8.790 | 5 | 0.118 |
| Probable menstrual cycle at scanning time, FL/LU/PM/UNK | 16/13/1/3 | | 15/8/5/3 | | 3.831 | 3 | 0.280 |
| Handedness, L/A/R | 0/1/32 | | 0/1/32 | | 0 | 1 | 1.000 |
| SD: standard deviation; df: degrees of freedom; S/M/SE/W: Single/Married or de facto/Separated or divorced/Widowed; PE/BU/T/L/H/ST/P/UN/Other: professional or executive/business/technician/labourer/housewife/student/pensioner/unemployed/other; FT/HTF/HTI: full time/half time fixed schedule/half time irregular; FL/LU/PM/UNK: follicular/luteal/postmenopausal/unknown (hysterectomy); L/A/R: left handed/ambidextrous/right handed  ^#^ Among those who work, Controls, N=27, Fibromyalgia, N=23  ^$^ The Socio-economic levels Questionnaire of the Mexican Association of Market Research and Public Opinion Agencies Questionnaire (AMAI NSE 8 × 7) indicates that A/B is the highest economic status category and E is the lowest | | | | | | | |
