## Supplementary material for "Cerebellar tissue properties alterations in fibromyalgia: a T1w/T2w ratio study": Table 2

| Table 2. Clinical characteristics |  | |  | |  |  |  |
| --- | --- | --- | --- | --- | --- | --- | --- |
|  | Controls (n=33) | | Fibromyalgia (N=33) | | Statistics | | |
|  | Mean | SD | Mean | SD | t-value/X^2^ | df | p-value |
| Time since diagnosis | - | - | 4.31 | 4.96 | - | - | - |
| Time since symptoms | - | - | 8.15 | 9.95 | - | - | - |
| Current medication received – Yes/No | - | | 30/3 | | - | - | - |
| Current medication received – Number of medications used daily 0/1/2/3/4/5 | - | | 11/11/4/4/2/1 | | - | - | - |
| Pain during interview (VAS)^$^ | 1.18 | 3.41 | 47.70 | 20.04 | **13.146** | **33.856** | **<0.001** |
| FIQ total score | - | - | 32.97 | 9.72 | - | - | - |
| McGill Pain questionnaire – sensory dimension | - | - | 39.13 | 14.57 | - | - | - |
| McGill Pain questionnaire – affective dimension | - | - | 7.31 | 3.24 | - | - | - |
| McGill Pain questionnaire – evaluative dimension | - | - | 3.31 | 1.80 | - | - | - |
| McGill Pain questionnaire – miscellaneous dimension | - | - | 8.88 | 4.13 | - | - | - |
| McGill Pain questionnaire – total score | - | - | 44.13 | 12.47 | - | - | - |
| Total widespread index (American College of Rheumatology criteria 2016)^$^ | .88 | 1.39 | 12.03 | 4.26 | **14.297** | **38.701** | **<0.001** |
| Total symptom severity score (American College of Rheumatology criteria 2016)^$^ | 1.42 | 1.42 | 8.39 | 2.42 | **14.269** | **51.549** | **<0.001** |
| HAMD total score | 1.21 | 1.93 | 15.58 | 6.37 | **12.388** | **37.834** | **<0.001** |
| HAMA total score | 2.18 | 2.43 | 21.55 | 6.83 | **15.338** | **39.964** | **<0.001** |
| SD: standard deviation; df: degrees of freedom; VAS: visual analogue scale; FIQ: fibromyalgia impact questionnaire; HAMD: Hamilton Depression Rating Scale; HAMA: Hamilton Anxiety Rating Scale  ^$^ Five healthy controls reported some pain during the interview, due to high physical activity in the previous days. None of the participant from the control group experienced any chronic pain condition | | | | | | | |
