## Supplementary material for "Cerebellar tissue properties alterations in fibromyalgia: a T1w/T2w ratio study": Table 3

| **Table 3. Results of the analyses using the severity of anxiety symptoms to mediate the group difference in T1w/T2w ratio values.** | | | | | | | | | | |
| --- | --- | --- | --- | --- | --- | --- | --- | --- | --- | --- |
| IV | DV | Mediator | Path | Estimate | Standardized estimate | se | z | p-value | 95%LLCI | 95%ULCI |
| ***Cerebellar Lobule VI*** | |  |  |  |  |  |  |  |  |  |
| Group | HAMA total |  | **a** | **19.342** | **0.886** | **1.262** | **15.324** | **<0.001** | **16.768** | **21.729** |
| HAMA total | Lobule VI |  | **b** | **-0.008** | **-0.468** | **0.003** | **-2.608** | **0.009** | **-0.014** | **-0.002** |
| Group | Lobule VI |  | **c’(total)** | **-0.206** | **-0.555** | **0.039** | **-5.315** | **<0.001** | **-0.278** | **-0.127** |
| Group | Lobule VI | HAMA total | c (direct) | -0.052 | -0.141 | 0.068 | -0.769 | 0.442 | -0.180 | 0.086 |
| Group | Lobule VI | HAMA total | **a*b (indirect)** | **-0.153** | **-0.414** | **0.061** | **-2.510** | **0.012** | **-0.275** | **-0.033** |
| ***Cerebellar Lobule VIII*** | |  |  |  |  |  |  |  |  |  |
| Group | HAMA total |  | **a** | **19.342** | **0.886** | **1.252** | **15.450** | **<0.001** | **16.843** | **21.753** |
| HAMA total | Lobule VIII |  | b | 0.001 | 0.051 | 0.004 | 0.240 | 0.810 | -0.006 | 0.008 |
| Group | Lobule VIII |  | **c’(total)** | **-0.181** | **-0.499** | **0.036** | **-4.997** | **<0.001** | **-0.249** | **-0.109** |
| Group | Lobule VIII | HAMA total | c (direct) | -0.197 | -0.544 | 0.081 | -2.423 | 0.015^#^ | -0.354 | -0.036 |
| Group | Lobule VIII | HAMA total | a*b (indirect) | 0.016 | 0.045 | 0.069 | 0.237 | 0.813 | -0.112 | 0.160 |
| IV: independent variable; DV: dependent variable, se: standard error; 95%LLCI: bootstrapped lower level 95% confidence interval; 95%ULCI: bootstrapped upper level 95% confidence interval; HAMA total: Hamilton Anxiety Rating Scale total score  Significant tests are in bold  ^#^ nominally significant did not survive the additional Bonferroni correction | | | | | | | | | | |
