## Supplementary material for "Cerebellar tissue properties alterations in fibromyalgia: a T1w/T2w ratio study": Table 4

| **Table 4. Results of the analyses using the severity of depressive symptoms to mediate the group difference in T1w/T2w ratio values.** | | | | | | | | | | |
| --- | --- | --- | --- | --- | --- | --- | --- | --- | --- | --- |
| IV | DV | Mediator | Path | Estimate | Standardized estimate | se | z | p-value | 95%LLCI | 95ULCI |
| ***Cerebellar Lobule VI*** | |  |  |  |  |  |  |  |  |  |
| Group | HAMD total |  | **a** | **14.347** | **0.839** | **1.157** | **12.405** | **<0.001** | **11.995** | **16.543** |
| HAMD total | Lobule VI |  | b | <0.001 | 0.010 | 0.004 | 0.060 | 0.952 | -0.007 | 0.008 |
| Group | Lobule VI |  | **c’(total)** | **-0.206** | **-0.555** | **0.038** | **-5.364** | **<0.001** | **-0.280** | **-0.128** |
| Group | Lobule VI | HAMD total | **c (direct)** | **-0.209** | **-0.564** | **0.058** | **-3.573** | **<0.001** | **-0.328** | **-0.094** |
| Group | Lobule VI | HAMD total | a*b (indirect) | 0.003 | 0.009 | 0.054 | 0.059 | 0.953 | -0.095 | 0.120 |
| ***Cerebellar Lobule VIII*** | |  |  |  |  |  |  |  |  |  |
| Group | HAMD total |  | **a** | **14.347** | **0.839** | **1.155** | **12.426** | **<0.001** | **11.950** | **16.493** |
| HAMD total | Lobule VIII |  | b | 0.003 | 0.121 | 0.004 | 0.674 | 0.500 | -0.005 | 0.010 |
| Group | Lobule VIII |  | **c’(total)** | **-0.181** | **-0.499** | **0.036** | **-5.017** | **<0.001** | **-0.248** | **-0.108** |
| Group | Lobule VIII | HAMD total | **c (direct)** | **-0.217** | **-0.601** | **0.071** | **-3.079** | **0.002** | **-0.361** | **-0.082** |
| Group | Lobule VIII | HAMD total | a*b (indirect) | 0.037 | 0.101 | 0.056 | 0.653 | 0.513 | -0.063 | 0.160 |
| IV: independent variable; DV: dependent variable, se: standard error; 95%LLCI: bootstrapped lower level 95% confidence interval; 95%ULCI: bootstrapped upper level 95% confidence interval; HAMD total: Hamilton Depression Rating Scale total score  Significant tests are in bold | | | | | | | | | | |
