## Supplementary Material for "Cerebellar tissue properties alterations in fibromyalgia: a T1w/T2w ratio study"

Yann Quidé^1,2,*^, Sylvia M. Gustin^1,2^

^1^ NeuroRecovery Research Hub, School of Psychology, The University of New South Wales (UNSW) Sydney, Sydney, NSW, Australia

^2^ Centre for Pain IMPACT, Neuroscience Research Australia, Randwick, NSW, Australia

***Corresponding author:**

Dr Yann Quidé, NeuroRecovery Research Hub, School of Psychology, Biological Sciences (Biolink) building, Level 1, UNSW Sydney, NSW, 2052, Australia.

| **Table S1. Results of the analyses using the T1w/T2w ratio values to mediate the group difference in severity of anxiety symptoms.** | | | | | | | | | | |
| --- | --- | --- | --- | --- | --- | --- | --- | --- | --- | --- |
| IV | DV | Mediator | Path | Estimate | Standardized estimate | se | z | p-value | 95%LLCI | 95%ULCI |
| ***Cerebellar Lobule VI*** | |  |  |  |  |  |  |  |  |  |
| Group | Lobule VI |  | **a** | **-0.206** | **-0.555** | **0.039** | **-5.342** | **<0.001** | **-0.279** | **-0.129** |
| Lobule VI | HAMA total |  | b | -8.520 | -0.145 | 3.591 | -2.372 | 0.018^#^ | -16.090 | -1.852 |
| Group | HAMA total |  | **c’(total)** | **19.342** | **0.886** | **1.247** | **15.508** | **<0.001** | **16.794** | **21.668** |
| Group | HAMA total | Lobule VI | **c (direct)** | **17.589** | **0.805** | **1.495** | **11.764** | **<0.001** | **14.501** | **20.341** |
| Group | HAMA total | Lobule VI | a*b (indirect) | 1.753 | 0.080 | 0.898 | 1.952 | 0.051 | 0.389 | 4.022 |
| ***Cerebellar Lobule VIII*** | |  |  |  |  |  |  |  |  |  |
| Group | Lobule VIII |  | **a** | **-0.181** | **-0.499** | **0.036** | **-5.072** | **<0.001** | **-0.250** | **-0.109** |
| Lobule VIII | HAMA total |  | b | 1.032 | 0.017 | 4.154 | 0.248 | 0.804 | -6.748 | 9.359 |
| Group | HAMA total |  | **c’(total)** | **19.342** | **0.886** | **1.248** | **15.503** | **<0.001** | **16.892** | **21.774** |
| Group | HAMA total | Lobule VIII | **c (direct)** | **19.528** | **0.894** | **1.541** | **12.673** | **<0.001** | **16.423** | **22.506** |
| Group | HAMA total | Lobule VIII | a*b (indirect) | -0.186 | -0.009 | 0.782 | -0.239 | 0.811 | -1.882 | 1.232 |
| IV: independent variable; DV: dependent variable, se: standard error; 95%LLCI: bootstrapped lower level 95% confidence interval; 95%ULCI: bootstrapped upper level 95% confidence interval; HAMA total: Hamilton Anxiety Rating Scale total score  Significant tests are in bold  ^#^ nominally significant did not survive the additional Bonferroni correction | | | | | | | | | | |

| **Table S2. Results of the analyses using the T1w/T2w ratio values to mediate the group difference in severity of depressive symptoms.** | | | | | | | | | | |
| --- | --- | --- | --- | --- | --- | --- | --- | --- | --- | --- |
| IV | DV | Mediator | Path | Estimate | Standardized estimate | se | z | p-value | 95%LLCI | 95%ULCI |
| ***Cerebellar Lobule VI*** | |  |  |  |  |  |  |  |  |  |
| Group | Lobule VI |  | **a** | **-0.206** | **-0.555** | **0.038** | **-5.381** | **<0.001** | **-0.280** | **-0.130** |
| Lobule VI | HAMD total |  | b | 0.204 | 0.004 | 3.281 | 0.062 | 0.950 | -6.553 | 6.418 |
| Group | HAMD total |  | **c’(total)** | **14.347** | **0.839** | **1.160** | **12.365** | **<0.001** | **11.996** | **16.518** |
| Group | HAMd total | Lobule VI | **c (direct)** | **14.389** | **0.842** | **1.436** | **10.021** | **<0.001** | **11.444** | **17.059** |
| Group | HAMD total | Lobule VI | a*b (indirect) | -0.042 | -0.042 | 0.699 | -0.060 | 0.952 | -1.273 | 1.526 |
| ***Cerebellar Lobule VIII*** | |  |  |  |  |  |  |  |  |  |
| Group | Lobule VIII |  | **a** | **-0.181** | **-0.499** | **0.036** | **-5.009** | **<0.001** | **-0.250** | **-0.107** |
| Lobule VIII | HAMD total |  | b | 2.652 | 0.056 | 3.855 | 0.688 | 0.491 | -4.395 | 10.850 |
| Group | HAMD total |  | **c’(total)** | **14.347** | **0.839** | **1.160** | **12.364** | **<0.001** | **12.011** | **16.586** |
| Group | HAMD total | Lobule VIII | **c (direct)** | **14.826** | **0.867** | **1.479** | **10.022** | **<0.001** | **11.855** | **17.680** |
| Group | HAMD total | Lobule VIII | a*b (indirect) | -0.479 | -0.028 | 0.739 | -0.648 | 0.517 | -2.258 | 0.725 |
| IV: independent variable; DV: dependent variable, se: standard error; 95%LLCI: bootstrapped lower level 95% confidence interval; 95%ULCI: bootstrapped upper level 95% confidence interval; HAMA total: Hamilton Anxiety Rating Scale total score  Significant tests are in bold | | | | | | | | | | |
